# A compound heterozygosity of two haplotypes in the TTN gene contributes to risk for Cleft Lip and Palate in a recessive manner

**DOI:** 10.1101/2025.01.15.24319654

**Authors:** Ved Prakash Singh, Subodh Kumar Singh, Akhtar Ali

## Abstract

Cleft lip and palate (CLP) are complex congenital anomalies with multifactorial etiology and significant genetic heterogeneity. This study aimed to elucidate the genetic basis of nonsyndromic CLP in an Indian family, wherein phenotypically normal parents had three affected offspring, including monozygotic twins, all presenting with bilateral complete CLP. Whole exome sequencing (WES) identified a compound heterozygous haplotype in the *TTN* gene shared by all affected siblings. The paternal haplotype comprised variants c.96140C>T (p.Thr32047Met), c.77412C>G (p.Phe25804Leu), c.2605A>T (p.Thr869Ser), and c.18663A>C (p.Glu6221Asp), while the maternal haplotype included variants c.107779G>A (p.Glu35927Lys), c.103147G>C (p.Glu34383Gln), c.63907G>A (p.Val21303Met), and c.26863A>G (p.Ile8955Val). In silico analyses predicted these variants to have deleterious effects on protein structure and function, consistent with a recessive mode of inheritance. Although *TTN* is primarily known for its role in muscle structure and function, emerging evidence implicates its involvement in craniofacial development. This study expands the phenotypic spectrum of *TTN*-associated disorders and suggests a novel role for *TTN* in nonsyndromic CLP. These findings underscore the importance of comprehensive genomic analyses in unraveling the molecular mechanisms underlying complex congenital anomalies.

## Introduction

Non-syndromic cleft lip with or without cleft palate, which affects most of the craniofacial region, is one of the most common birth abnormalities in humans (1). The facial prominences fail to properly fuse in a coordinated way during embryogenesis, leading to these developmental abnormalities (2). Approximately 70% of orofacial clefts (OFCs) are nonsyndromic (nsOFCs), which includes this particular birth abnormality as one of its subtypes. Nonsyndromic cleft palate only (nsCPO) is one of the other subtypes of nsOFCs. 1/700 live births are said to represent the aggregate worldwide prevalence of OFCs (3). Malocclusion, eating issues, communication deficits, and aesthetic issues are among the associated disabilities brought on by these deformities. According to studies, those with nsCL/P had a significantly higher overall death rate (4). Families of those impacted by the disease have expressed severe financial, psychological, and social hardships as well as frequent stigmatization (5). For effective care, a group of experts must handle difficulties with speech, dentistry, and psychology in addition to performing surgical repair of the problem (6). Orofacial clefts have a complicated aetiology that is related to many embryological origins and developmental stages (7,8). A cleft lip results from the failure of the primary palate, whereas a cleft palate is the result of the secondary palate’s failure to form (9). Due to the distinct developmental origins of the primary and secondary palates, Cleft palate only (CPO) is considered separate from CL and CLP, although shared aetiological factors are still addressed (10). Subclinical phenotypes of cleft palate include submucous cleft palate and bifid uvula. While main palate construction is completed by weeks seven or eight, secondary palate formation begins around week seven and lasts until week twelve (11). The interplay between genetic changes and environmental influences might be regarded as the etiology of orofacial clefts (12,13). Orofacial clefts have been revealed to be associated with an abundance of potential genes and loci (14,15,16). The existence of orofacial clefts may also be influenced by environmental variables like as smoking, drinking alcohol, deficiency in certain vitamins and minerals, age of parents, exposure to pollutants in the environment, and socioeconomic level (17,18,19,20). There is additional evidence that the age of the parents increases the chance of orofacial cleft (21). The role of genetics in the genesis of clefts has been widely reported; the estimated heritability rate for clefts ranges from 50 to 80% (22). Just over 40% of cleft instances have been directly related to genetic risk factors, even though all cases tend to exhibit familial tendencies (23). The occurrence of orofacial clefts in a family suggests that this condition follows heredity. A study found that the risk of recurrence is 32 times higher for first-degree relatives of individuals with CL/P than for those without a positive family history (24). This study found that first-degree relatives had a 56-fold increased relative risk of CP recurrence compared to the population-based control group. (24). The third-degree relative risks are not substantially different from the population-based control, which suggests that the relative risk of clefts in affected families decreases as the degree of segregation increases (24). Concordance rates of clefts in monozygotic twins (40–60%) were found to be greater in twin studies than in dizygotic twins (3-5%) (25). A parent with nsCL/P has a 3.2% chance of producing an afflicted offspring, according to other studies (24). The probability of the problem in a subsequent birth rises to 15.8% after the delivery of an afflicted kid to such a parent (24). On the other hand, there is a 4.4% chance that unaffected parents of an affected kid will also give birth to an affected child (24). When these are considered collectively, a significant genetic factor in the etiopathogenesis of clefts becomes apparent. Whole-exome sequencing (WES) has been used to find minor insertions, deletions, and pathogenic single nucleotide polymorphisms (SNPs) associated with nsCL/P in different ethnic groups (26,27,28,29,30,31). Using whole exome sequencing on 46 multiplex OFC families, Basha et al. discovered that 10% of cases had “likely pathogenic” (LP) mutations, mostly in genes that cause AD OFC syndromes (32).

## 2. Materials and Methods

### 2.1 Ethical approval and informed consent

This study is approved by the Ethics Review Committee of the Institute of Science, Banaras Hindu University (BHU). The parents provided written informed consent for the study to collect and use the clinical data, images, and samples. Every procedure involving human subjects was carried out by the institutional or Indian Council of Medical Research guidelines for ethical standards.

### 2.2 Sample collection

An Indian family was recruited by co-author Dr. S.K. Singh. Parents were normal with the first proband a male child having “Bilateral complete Cleft lip and cleft palate” and the second proband was a monozygotic twin girl with “Bilateral complete Cleft lip and cleft palate”. Pedigree is shown in figure 1. Photographs and peripheral blood samples were taken from all family members. The comprehensive clinical evaluation of probands was done to find any other anomalies and no other anomalies were found which categorizes it as a non-syndromic cleft lip and palate case.

**Figure 1:**
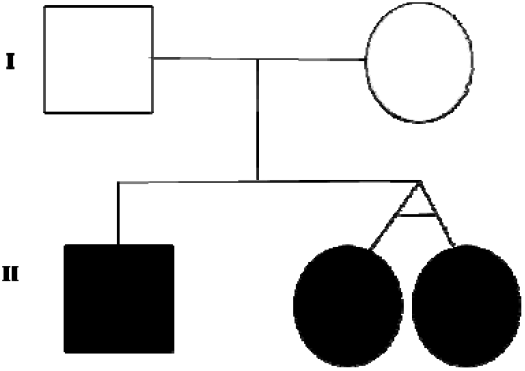

### 2.3 DNA Extraction and Whole Exome Sequencing

The salting-out approach extracted DNA from peripheral blood (Miller et al., 1988). DNA was quantified using the spectrophotometer (NanoDrop ND-2000, Thermo Fisher Scientific, USA).

### 2.4 Library Preparation

The Whole Exome libraries were constructed using MGIEasy Exome FS Library Prep Kit. The genomic DNA of 200 ng was subjected to fragmentation and purified using MGIEasy DNA Clean beads. The purified fragmented DNA was end-repaired and dA-tailed. Further, adapter ligation was performed to attach sequencing technology-specific adapters containing the index. MGIEasy DNA Clean beads were utilized to purify the adapter-ligated DNA. The adapter ligated and indexed DNA was PCR amplified for 8 cycles followed by purification using MGIEasy DNA Clean beads. Quality check of the library was performed using Qubit Fluorometer (Thermofisher Tech.) and Agilent Bio-analyzer. Further, the libraries were pooled in equal concentration to set up a hybridization reaction for target capture using MGIEasy Exome Capture V5 Probes for 24 hours. Target capture was performed using Streptavidin beads, and then the on-bead library was PCR amplified for 12 cycles. A quality check of the targeted library was performed.

### 2.5 Sequencing

The dsDNA libraries were subjected to ssDNA circularization and DNB formation as per loading requirements. The libraries were sequenced on G400 MGI sequencer as per the manufacturer’s protocol.

### 2.6 Data Analysis

#### 2.6.1 Quality Check and Alignment

A post-sequencing quality check was performed using FASTP and FASTQC software. Adapters and low-quality reads were removed during the quality assessment. For more than 92% of the targeted regions, the average depth of coverage was 100X. Afterward, these clean reads were aligned with human genome version GRCh38 using Burrows-Wheeler Aligner (BWA) (http://bio-bwa.sourceforge.net/) software.

#### 2.6.2 Variant Calling

The GATK Haplotype Caller algorithm (https://www.broadinstitute.org/gatk/) was utilized for base quality recalibration and variant calling. Sequence Alignment/Map tools (SAMtools) identified the presence of single nucleotide polymorphisms (SNPs) and insertions/deletions.

#### 2.6.3 Variant Annotation

Variant annotation was performed using various open-source software and databases. Clinically relevant mutations were annotated using published literature variants and disease databases, i.e., ClinVar, OMIM, and HPO. The 1000Genome (https://www.internationalgenome.org/), gnomAD (https://gnomad.broadinstitute.org/), and dbSNP (https://www.ncbi.nlm.nih.gov/snp/) were used for annotation of variants minor allele frequency. Functional prediction of all potential non-synonymous variants was done using dbNSFP.

#### 2.6.4 Variant Filtering

For missense, nonsense, splicesite, stop loss, start codon change, frame-shift, and in-frame indels, variant filtering was carried out using MAF<1%. Various other parameters such as depth quality>=20, genotype quality >30 etc. were taken care during variant f i ltration. All synonymous variants were removed from the analysis.

Pathogenic prediction scores were calculated for non-synonymous variants using Polyphen2 (http://genetics.bwh.harvard.edu/pph2), SIFT (http://sift.jcvi.org), MutationTaster (http://www.mutationtaster.org/), and Mutation Assessor (http://mutationassessor.org) in order to assess the effect of amino acid substitution on protein structure and function. To draw attention to particular phenotypes, such as HP:0410030 (cleft lip), HP:0000175 (cleft palate), HP:0002006 (facial cleft), HP:0000202 (oral cleft), HP:0100333 (unilateral cleft lip), and HP:0100334 (unilateral cleft palate), we used the Germline Managed Variant List filter.

Standard nomenclature, such as pathogenic, likely pathogenic, unknown significance, likely benign, and benign variants, was used to characterize variants found in genes, according to the American College of Medical Genetics and Genomics (ACMG) guidelines.

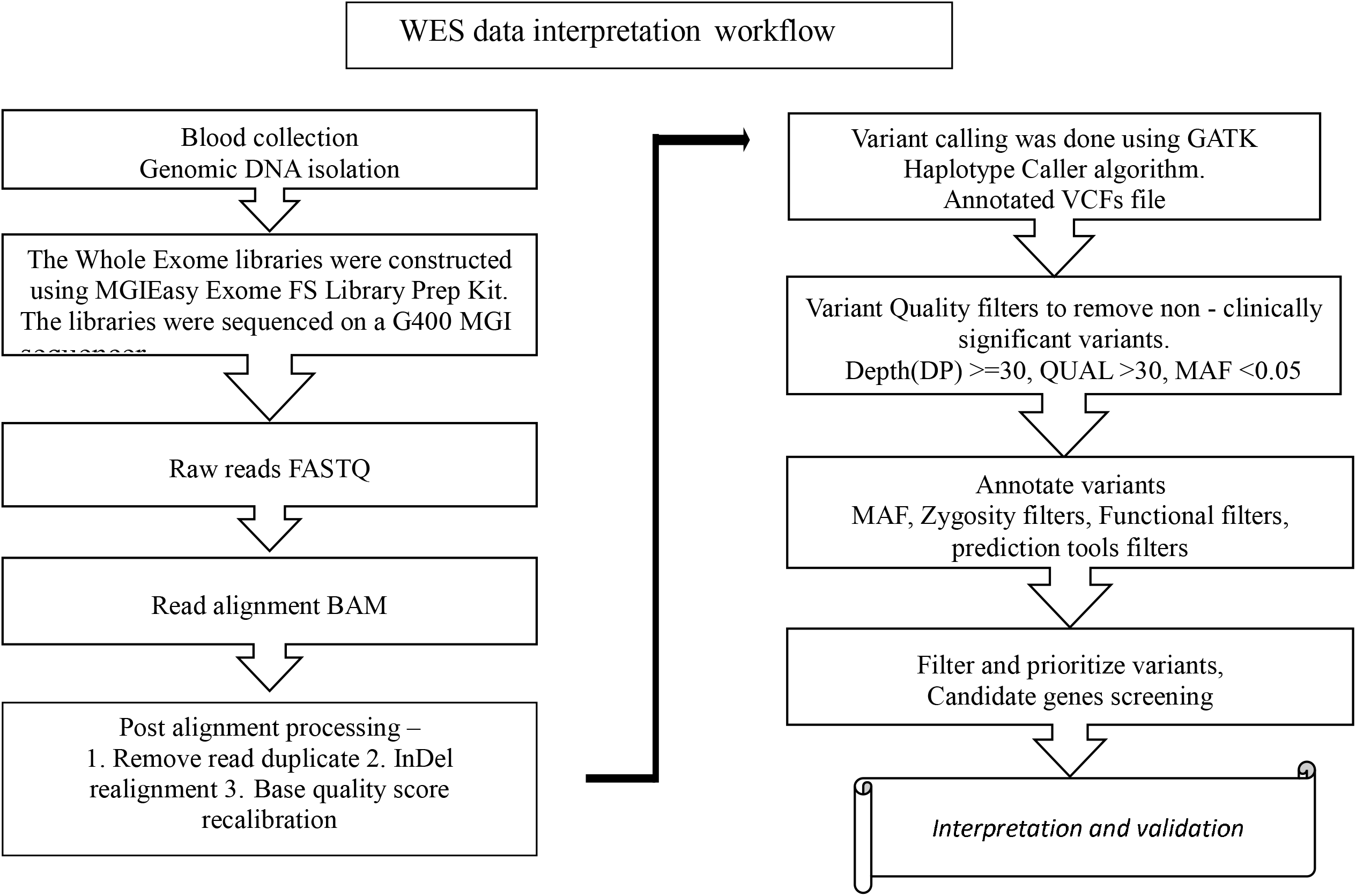

## 3. Results

Whole exome sequencing was done on the G400 MGI sequencer platform. In the framework of a preliminary analysis conducted, we first detected 41235 variants in father, 39077 variants in mother,28154 variants in son, and 39638 variants in daughter-1 & 39085 variants in daughter-2 across the whole genome.

We focused on the selected candidate gene of cleft lip and palate, the gene was taken from a literature review, NCBI database, OMIM, and Genecard database. 172 genes were listed for analysis, we focused on the pattern of inheritance of coding variants of these genes.

We found compound heterozygous haplotypes of the *TTN* gene in all the affected sibs. In father c.96140C>T, c.77412C>G, c.2605A>T and c.18663A>C missense variants are present in the heterozygous condition which changed the amino acid p.Thr32047Met, p.Phe25804Leu, p.Thr869Ser and p.Glu6221Asp respectively.

In mother c.107779G>A, c.103147G>C, c.63907G>A and c.26863A>G missense variants are present in the heterozygous condition which changed the amino acid p.Glu35927Lys, p.Glu34383Gln, p.Val21303Met and p.Ile8955Val respectively (Table:1).

**Table 1:** Quality Parameters.

| Samples I.D. | Data (GB) | Mean Depth | Read Length | No of Reads | Q30 (%) |
| --- | --- | --- | --- | --- | --- |
| CVF-01 | 13.2 | 105 | 100 | 132007398 | 83.24 |
| CVM-02 | 13 | 103 | 100 | 129522826 | 83.58 |
| CVS-03 | 12.2 | 99 | 100 | 122400274 | 83.33 |
| CVIK-04 | 12 | 97 | 100 | 120456886 | 83.91 |
| CVIL-05 | 12.6 | 102 | 100 | 125923148 | 82.59 |

**Table 1:** Details of the *TTN* gene variants identified in the family members.

| Chromosome:<br>Position (Hg38) | rs ID | Exon | Nucleotide<br>change | Genotype | Amino acid<br>change |
| --- | --- | --- | --- | --- | --- |
| 2:178527209 | rs751773148 | 363 | c.107779G>A | CT | p.Glu35927Lys |
| 2:178533468 | rs148525155 | 358 | c.103147G>C | CG | p.Glu34383Gln |
| 2:178544004 | rs375640847 | 346 | c.96140C>T | GA | p.Thr32047Met |
| 2:178568720 | rs536177370 | 326 | c.77412C>G | GC | p.Phe25804Leu |
| 2:178587304 | rs372812312 | 307 | c.63907G>A | CT | p.Val21303Met |
| 2:178713271 | rs72648994 | 93 | c.26863A>G | TC | p.Ile8955Val |
| 2:178729493 | rs369544339 | 64 | c.18663A>C | TG | p.Glu6221Asp |
| 2:178784240 | rs370962244 | 16 | c.2605A>T | TA | p.Thr869Ser |

All affected siblings, the first male child (Son)and the second monozygotic female twin (Daughter-1 & Daughter-2) share both parental haplotypes (Table:2). This pattern of inheritance shown in figure 2.

**Table 2:** Showing patter of inheritance in siblings from parents.

| GENE | Father | Mother | Son | Daughter-1 | Daughter-2 |
| --- | --- | --- | --- | --- | --- |
| ***TTN***<br>**(2q31.2)** | **c.65348A>T** | normal | normal | normal | normal |
|  | **c.55616T>C** | normal | normal | normal | normal |
|  | **c.96140C>T** | normal | **c.96140C>T** | **c.96140C>T** | **c.96140C>T** |
|  | **c.77412C>G** | normal | **c.77412C>G** | **c.77412C>G** | **c.77412C>G** |
|  | **c.2605A>T** | normal | **c.2605A>T** | **c.2605A>T** | **c.2605A>T** |
|  | **c.18663A>C** | normal | **c.18663A>C** | **c.18663A>C** | **c.18663A>C** |
|  | normal | **c.107779G>A** | **c.107779G>A** | **c.107779G>A** | **c.107779G>A** |
|  | normal | **c.103147G>C** | **c.103147G>C** | **c.103147G>C** | **c.103147G>C** |
|  | normal | **c.63907G>A** | **c.63907G>A** | **c.63907G>A** | **c.63907G>A** |
|  | normal | **c.26863A>G** | **c.26863A>G** | **c.26863A>G** | **c.26863A>G** |
|  | normal | **c.24880A>G** | normal | normal | normal |
|  | normal | **c.\*99dupA** | normal | normal | normal |

**Table 3:**
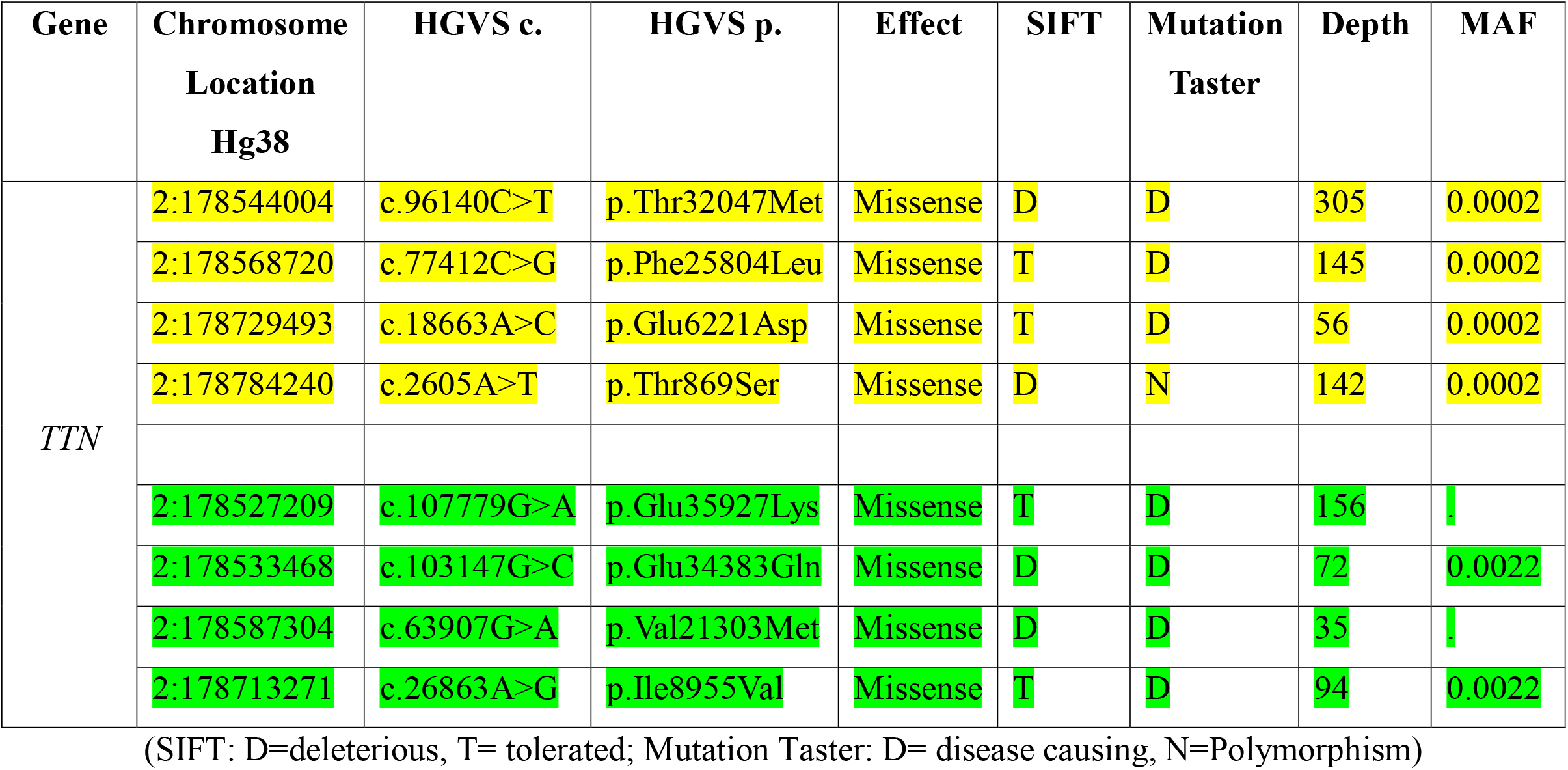
(Bioinformatics analysis of variants)

**Figure 2:**
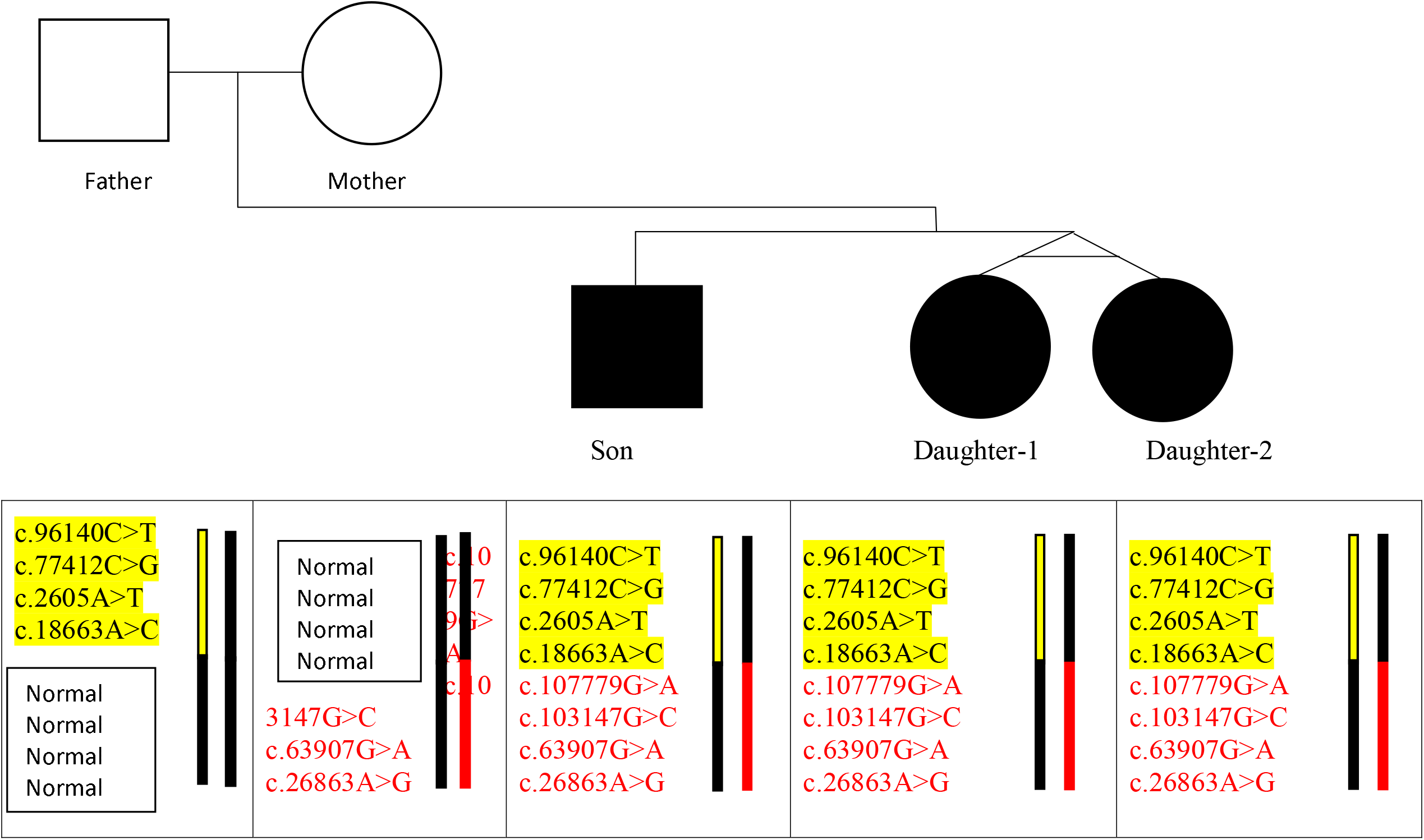
The inheritance of the two *TTN* haplotypes one from father and another from mother. All the affected children Further bioinformatics analysis shows the effect of identified variants (Table:3).

## Discussion

Cleft lip and palate is a multifactorial birth disorder. Genetic heterogeneity in the manifestation of CLP is complex in itself. Some genes like *CDH1, TP63, IRF6, NECTIN1*, and *SMX1* are well-known candidate genes for CLP. Mutation in these genes are associated with cleft lip and palate. Several WES studies have been done and identified known as well as novel variants in CLP patients. These variants are found in homozygous recessive or heterozygous conditions. There is the possibility that variants or variants present in a healthy person may act as a causal factor in the affected person if such variants are present in a compound. So to find such results whole exome sequencing of the family is needed.

We performed WES of a family in which the parents were healthy but there all the children were cleft lip and palate patients. The analysis found a compound heterozygous haplotype of the *TTN* gene present in all affected siblings. All affected siblings inherited the compound heterozygous haplotype of the *TTN* gene, suggesting that these genetic variants may play a significant role in the etiology of their cleft lip and palate. *TTN* gene comprises 364 exons located on chromosome 2q31 and is the largest protein-coding gene (33). It plays a crucial role in the structure and function of striated muscles. It contributes to muscle elasticity and integrity, which are vital during embryonic development (34). Mutation in *TTN* has been associated with congenital myopathies (35).

Studies suggest that titin plays a role in important developmental processes, including those that influence the development of facial features. Research involving mutant mice showed that alterations in the TTN gene increased apoptosis of cells in the frontonasal process, a critical tissue for lip and palate formation (36). This implies that titin function abnormalities could be a direct cause of the mechanisms behind CLP.is implies that titin function abnormalities could be a cause of the mechanisms behind CLP. Cleft palate has also been reported in some individuals with cardiomyopathy having *TTN* mutation (37). In a recent study, De novo mutations (DNMs) in the *TTN* gene have been found through whole-genome sequencing to be linked to nonsyndromic cleft lip with or without cleft palate (38). Developmental abnormalities may result from these mutations, which can interfere with proper titin activity.

Importantly, the inheritance of these particular variants in compound heterozygosity is limited to the affected siblings, indicating a recessive mode of inheritance involving *TTN* in the etiology of cleft lip and palate in this family, even though specific instances of compound heterozygous variants in *TTN* leading to CLP are uncommon.

## Conclusion

The identification of compound heterozygous variants in the *TTN* gene in all affected siblings suggest that these genetic alterations contribute to the development of CLP in this family. Both parents have several amino acid-changing variations, which suggests that although they are phenotypically normal, they have genetic predispositions that may cause their children to exhibit CLP. The TTN gene is one of the critical genes associated with the development of numerous myopathies, and its link with clefting disorders further broadens its clinical spectrum. This case highlights the necessity for detailed genetic analysis in complex congenital abnormalities like cleft lip and palate, especially when there are normal phenotypes in parents but affected children. This case points towards the necessity of the involvement of whole genetic analysis in characterizing any complex congenital disorders especially cleft lip and palate by involving families with a normal phenotype but an abnormal child.

## Data Availability

All data produced in the present work are contained in the manuscript.

